# Generating Biologically Relevant Subtypes of Autism Spectrum Disorder with differential responses to Acute Oxytocin Administration in a Randomized Trial using Random Forest Models and K-means Clustering

**DOI:** 10.64898/2026.02.10.26346006

**Authors:** Christopher D. Vento, Jolee Hatfield-King, Kaundinya Gopinath, Shota Nishitani, Michael Morrier, Opal Ousley, Joseph F. Cubells, Larry J. Young, Elissar Andari

## Abstract

Autism Spectrum Disorder (ASD) is a heterogenous condition that has no biologically relevant subtypes yet. Here, we utilized a multidimensional approach considering social deficits in ASD alongside negative valence and empathy dysfunction to distinguish ASD from Neurotypicals (NT) and to generate ASD subtypes using machine learning approaches. 114 subjects were analyzed, with 70 being NT and 44 ASD, all male with an IQ greater than 70, with 5 domains of personality (NEO-PI-r) and Reading the Mind the Eyes Test (RMET) scores included in the main classifier. We then used a multitude of behavioral (such as IQ, Broader Autism Phenotype, Autism Quotient, Interpersonal Reactivity Index) and clinical measures such as Autism Diagnostic Interview-Revised (ADI-R) alongside biological methods including DNA methylation of *OXTR* gene and resting-state functional connectivity (rsFC) to validate the putative subtypes. 30 ASD who received IN-OXT in a randomized, placebo-controlled, within-subject design and 17 new NT were part of the rs-FC analysis. A random forest tree algorithm was used to classify NT and ASD and Shapley Additive Explanation Values were used to describe the model and to cluster ASD subtypes using K-Means clustering. Three subtypes were generated with two of them being highly distinctive in behavioral and brain functional traits. One subtype named NASA (or Negative Affect and Social Aloofness) was characterized by high Neuroticism and Low warmth alongside lower rsFC between networks involved in social cognition, self-awareness, and sensory processing, such as Superior Temporal Sulcus and Sensorimotor Network; or ACC/Insula with visual cortex, Posterior Cingulate Cortex and visual cortex. The second subtype NADR (Neurocognitive and Affect Dysregulation with Resistance to Change) was characterized by higher DNA methylation of *OXTR*, hyperconnectivity between default mode network, reward areas and inferior frontal and fusiform networks. NADR has more cognitive difficulties and higher ADI-R scores as well as higher Neuroticism, higher personal distress, higher rigidity and lower openness. In a mixed model analysis, we found that IN-OXT in a dose dependent manner impacted NASA subtype by modulating rsFC between PCC and cerebellum and between Brainstem/Cerebellum and Parietal cortex to probably enhance social cognition and to reduce negative valence in this subtype.

## Introduction

It has been a great challenge to find biomarkers and effective treatments for autism spectrum disorder (ASD) given the striking intrinsic phenotypical and biological heterogeneity. With the prevalence on the rise with 1 in 31 children are diagnosed with ASD according to recent records [1], generating ASD subtypes that are characterized by distinctive features and dysfunctions and underlying biological mechanisms is fundamental to make progress in finding adequate intervention or care.

In the past, researchers have applied clustering, latent profile, mixture modeling, and graph-based approaches to behavioral, and biological data to parse ASD into more homogeneous subtypes [2–17]. While these studies consistently demonstrate that ASD is not a singular condition, there is still lack of consensus on what subtypes are biologically relevant and on whether some of them respond to treatment.

Early and ongoing efforts to subtype ASD using behavioral measures have demonstrated that subgroups can be generated based on levels of dysfunction in social functioning, sensory processing, or adaptive behavior. Studies using standardized tools identified subgroups spanning between severe, moderate, and relatively adaptive characteristics [14,18]. These subgroups differ in core autism features and in cognitive ability, emotional and behavioral problems, and quality of life, highlighting the multidimensional nature of ASD. Studies focusing on sensory processing grouped children with ASD into over-responsive, under-responsive, sensory-seeking, and mixed subtypes, that relate differently to emotion dysregulation, anxiety, ADHD symptoms, and everyday functioning [19,20]. *Despite their strengths, behavioral-based subtyping studies are mostly focused on unidimensional features (social processes or sensory processing or other), often lead to severity-based classification (severe versus mild) and not qualitative differences, yield subtypes that vary across studies, and limit insight into underlying neurobiological mechanisms, restricting their utility for precision categorization and intervention*.

Studies incorporating neuroimaging and other biological measures often used large datasets such as *Autism Brain Imaging Data Exchange* (ABIDE) [21]. ASD subtypes were generated based on patterns of cortical thickness, and gray and white matter volume [22,23], or on functional hyperconnectivity or hypoconnectivity, involving sensorimotor and subcortical networks with associated differences in symptom severity, cognition, and adaptive functioning, which suggest biological relevance [24,25]. Studies that combine behavioral and neural measures provide a more inclusive route for identifying meaningful subtypes of ASD. For instance, Zabihi et al. (2020) showed that combining cortical thickness with behavioral data provides subgroups that differ in cognitive ability, social functioning, and severity of restricted interests and repetitive behaviors [8]. Collectively, there is a meaningful heterogeneity that can be detected with behavioral studies lacking biologic understanding, and biological studies lacking clinically distinguishable phenotypes. Importantly, there is a lack of replicable subtypes and lack of validity in terms of personalized treatments based on subtypes.

*Here, we propose to use multimodal (behavior, neuroimaging, epigenetics, clinical features) and multidimensional approaches (social behavior, negative state valence including anxiety and depression, empathy, cognition) to conduct deep phenotyping on domains of function that are related to core deficits of ASD and comorbidities such as Negative Valence. Prevalence of associated anxiety in ASD is as high as 60% based on recent estimates [26,27]. The goal is to examine if there are subtypes of autism that are more responsive to drug treatment that we used in our previous studies, oxytocin [28]*.

<u>We hypothesize that the use of the following comprehensive measures can help uncover biologically relevant subtypes that account for the heterogeneity of symptoms in ASD and that can be targeted by selective treatment based on shared mechanisms of action.</u>

We used *Random Forest Models* and *Shapley Additive exPlanations* (SHAP) to first differentiate ASD and NT from each other. We then clustered the individual SHAP with *K-Means* to generate various subtypes within ASD [29,30]. In the classifier, we included a comprehensive personality measure (NEO-PI-r) that includes domains of Extraversion, Neuroticism and other; and the Reading the Mind in the Eyes Test (RMET) that consists of identifying emotional states from the eye region as a measure of empathy that is also relevant to ASD physiopathology as well [31,32]. Biological validation measures include clinical diagnostic criteria, resting-state functional connectivity (rs-FC), and DNA methylation of the oxytocin receptor gene (*OXTR*) that was previously found hypermethylated in ASD among other measures that were not part of the initial classifier [33].

Furthermore, we examined whether any of the generated subtypes respond better to a particular treatment that is part of the previous Autism Oxytocin Brain project (https://www.clinicaltrials.gov/study/NCT03033784). Participants received several doses of intranasal oxytocin (IN-OXT) 40 minutes prior to going through a resting-state functional connectivity fMRI scan at the Emory University Hospital. Here, we are examining whether any of the subtypes generated from this study respond more effectively to acute intake of IN-OXT using brain rsFC as target engagement for the drug. IN-OXT has shown some promising results on enhancing social skills in ASD and some inconsistencies in long-term clinical trials [34–40]. Studying if there is a subtype that would respond best to IN-OXT can partially address these discrepancies.

## Methods and Materials

### Subjects

Subjects were recruited from Emory University as part of the Autism Oxytocin Brain Project. All subjects were adult males and between 18 and 45 years old with an Intelligence Quotient greater than 70. We initially recruited 74 NT individuals and 50 individuals with ASD to conduct a series of behavioral tasks and saliva collection for DNA methylation analysis of the oxytocin receptor gene following procedures and obtaining participants’ informed consent as approved by the Emory Institutional Review Board (IRB). 10 ASD subjects were excluded because of eligibility criteria which led to a final N of 40 for ASD sample size for behavioral data collection. One subject withdrew from the study and therefore the total number of ASD was 39 for the behavioral analysis. Three NT individuals did not complete the NEO-PI-r which led to 71 as the total number of NT for data analysis.

Only 30 adults with ASD completed the clinical trial with oxytocin and fMRI (resting state as one of the tasks) as approved by the Emory IRB. We also included a new set of NT (17 male controls for IQ and for age and race) for the fMRI results (see Supplementary file and previous publication) [28]. 2 ASD subjects did not have data for the NEO-PI-r personality inventory and therefore were excluded from the analysis. This gives us a sample of 28 total ASD for the neuroimaging analysis. The study was approved by the Emory IRB. Participants started testing procedures after reading and signing appropriate written-informed consent documents following discussion with staff of any questions or concerns.

### Behavioral tests, surveys, and clinical assessments

All subjects were tested with the written self-report personality inventory NEO-PI-r [32], the Reading the Mind in the Eyes or RMET task [41,42], the Symptom Checklist or SCL-90-R [43,44], the Wechsler Adult Intelligence test (WASI-II) [45,46], the Social Responsiveness Scale or SRS-2 [47,48], the Broader Autism Phenotype Quotient or BAPQ [49], and the Autism Quotient AQ[50]. Those who were selected for the clinical trial of iN-OXT also completed the Interpersonal Reactivity Index or IRI as a measure for empathy which includes measures of perspective taking, fantasy, empathic concern, and personal distress [51].

Finally, the *Autism Diagnostic Observation Schedule, 2^nd^ edition* (ADOS-2) and *Autism Diagnostic Interview-Revised (ADI-R)* tests were used to confirm ASD diagnosis. These tests were used to validate the ASD subtypes and to detect any measurable differences between them [52,53].

### Methylation Methods

Please refer to Supplementary file for details and to Andari et al., 2020 for the detailed methods of epigenetic methylation analysis of *OXTR* gene from salivary samples [54].

### Resting State fMRI acquisition, pre-processing and analysis

See Supplementary file and Andari et al., 2025 for detailed description of resting-state functional independent component analysis [28].

### Intranasal oxytocin administration

The study consisted of a randomized, placebo-controlled, double-blind, within-subjects design in which each participant received 48 IU, 24 IU, and 8 IU of Syntocinon Spray (Novartis) (three puffs per nostril) or placebo 40 min before resting-state task, across 4 randomized visits. Patients were instructed to keep their eyes open and look at a fixation cross inside the MRI scanner 40 minutes after taking the nasal spray (See Andari et al., 2025 for more details) [28].

### Machine Learning and Statistical Methods

#### Classifying ASD and NT

RMET and NEO-PI-r scores were included for both groups. All machine learning and statistical analysis were conducted in Python [55]. Missing data points were imputed using the missForest from the missingpy package [56]. A hyper parameterized gridsearch using SciKit-Learn was then conducted to find the most optimal parameters for a Random Forest Tree Model comparing the ASD and NT. Accuracy, Specificity, ROC, and AUC were calculated for the Random Forest Tree model [57]. Shapley Additive exPlanations values (SHAP) were calculated using the SHAP package from the Random Forest Tree Model to assess the relative contributions of each feature [29,30].

### Clustering ASD

SHAP values of the individual subjects were subsequently calculated. The log odds for diagnoses of ASD and NT were then calculated for each SHAP frame and then normalized [29,30]. NT were then excluded from analyses focusing on the ASD group alone. The ASD Normalized SHAP and Log Odds were visualized together using Principal Component Analysis and clustered using K-Means Clustering from the Sci-Kit Learn Package. The optimal number of clusters was calculated by calculating the inertia values for the individual in number of subtypes [57]. Once the final model was selected, SPSS version 30 was used to calculate one-way ANOVAs with type III sum of squares and eta-squared effect size used to compare subtypes and NT. Post-Hoc T-Tests with Bonferroni adjustment followed this analysis [58].

### Statistical Analysis

Statistical analysis was done in SPSS version 30 [58]. All data were assessed for outliers by transforming standardized Z–scores of all values for those data and removing individual datapoints that had a z-score less than –3 or greater than 3.

A one-way analysis of variance was used to individually examine the differences in the NEO-PI-r main domains and facets, RMET, BAPQ, SCL-90R, Autism Quotient, Intelligence Quotient, IRI, methylation, and resting state neuroimaging, including the 3 different subtypes of ASD and NT. Post-Hoc T-Tests with Bonferroni adjusted P-Value were then used to examine average between-group differences.

A one-way analysis of variance was used to individually examine the differences in ADOS-2, ADI-R, and IQ across 3 levels including the 3 different subtypes of ASD. Post-Hoc T-Tests with Bonferroni adjusted P-Value were then used to examine average between-group differences.

To test whether Subtypes of ASD were affected differently by OXT treatment, we conducted a linear mixed mode analysis using SPSS version 30 [58]. Individualized functional brain networks were used as dependent factors, the different doses of IN-OXT treatment, the 3 subtypes, and the dose-subtype interactions between were used as fixed factors and the subject ID was used as a random factor, with age in years as a covariate. Type III sum of squares was used to estimate the main effects and the interactions. The variance component of the mixed model was calculated using restricted maximum likelihood. Finally, the marginal means for each different combination of subtypes and dosages were compared using t-tests with Bonferroni correction to compare differences in brain activity changes across dosages among the individual subtypes. We only reported results that are significantly different from placebo.

## Results

### 1- Machine learning models – Classifying ASD and NT and contributor factors

Hyperparamaterized features of the Random Forest Tree identified the following optimal model with 1131 estimators, 0.37 max features, 4 minimal sample leaves, and balanced subsample class weights. The final model has an average accuracy of 0.78 ± 0.09 with an AUC of 0.82 ± 0.14, with a specificity of 0.87 ± 0.09, and with a precision of 0.82 ± 0.1, F1 0.84 ± 0.07 differentiating ASD from neurotypicals (NT) (see Figure 1A). (See Supplementary Result for additional models tested).

**Figure 1:**
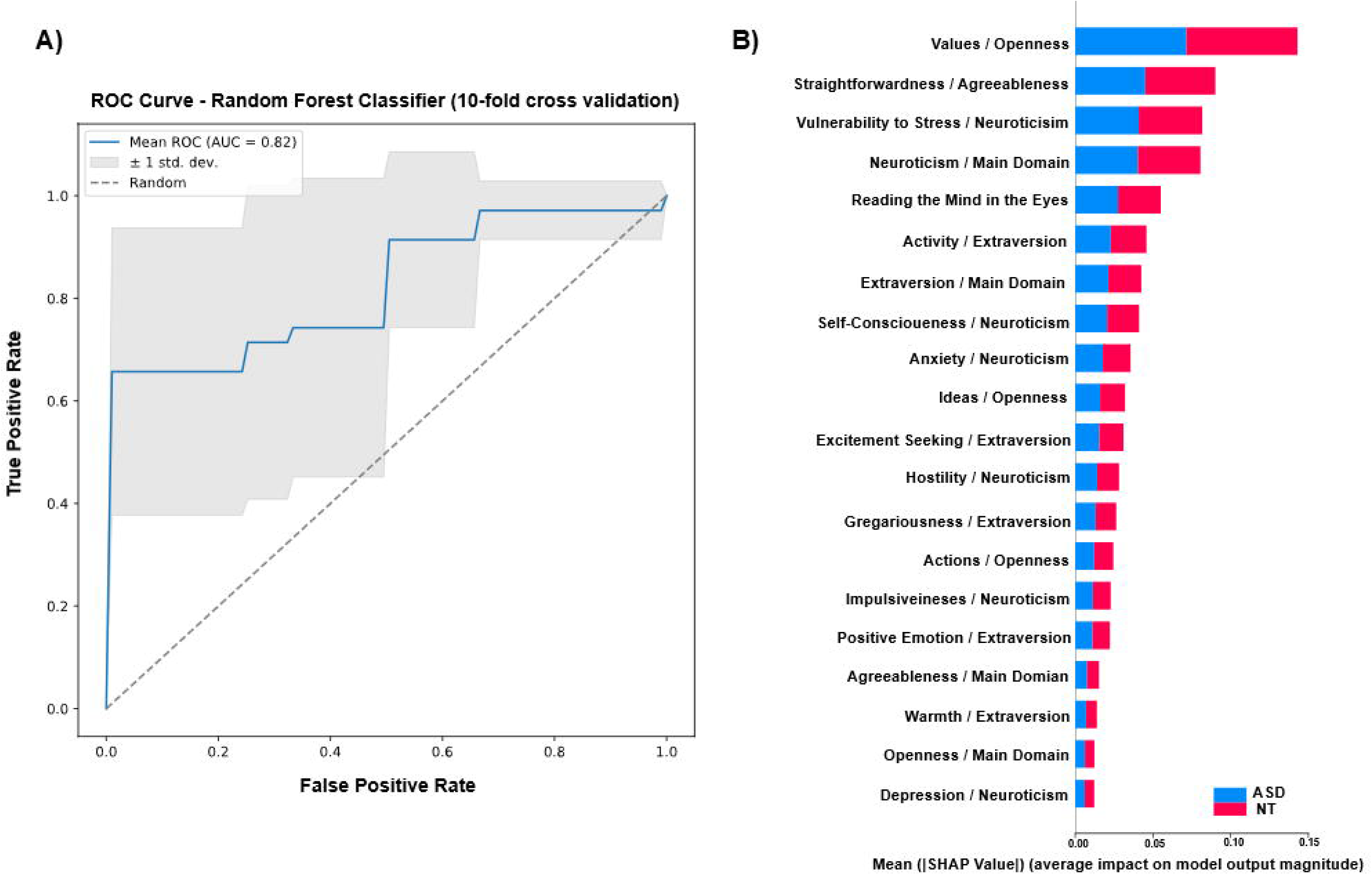
(A) ROC Curve Curve from Random Forest Classifier with 10-fold cross-validation with False Positive Rate on the X-axis and the True Positive Rate on the y-axis with the average performance of the model been shown in the blue line the gray bars being plus or minus one standard deviation with a calculated AUC of 0.81. The dialog line represents the model’s performance due to chance. Based on this figure we can infer that the model does significantly better due to chance with the model doing better than chance. Figure (B) SHAP values that show the individual features contributions with the average impact on the model output magnitude being shown for the top features. The total bar length represents the overall features performance, while the blue and red colors show the individual contributions towards classifying a subject as ASD or NT.

In the current study, *Shapley Additive exPlanations values* (SHAP) analysis indicated that NEO-PI-r domains of **Openness, Neuroticism, and Extraversion** are among the strongest contributor factors differentiating ASD from NT in the best-fitting model (Fig 1A). Several facets of **Neuroticism** are among the top contributors such as *Vulnerability to Stress*, *Self-Consciousness, Anxiety, and Hostility/Anger*. For the facets of **Extraversion**, *Activity* and *Excitement seeking* are contributors as well. For the facets of **Openness**, *Values* is the top contributor to the classifier alongside *Ideas* that is among the top 10 contributors (Figure 1B). *Straightforwardness* from the domain of **Agreeableness** was the second top contributor. **RMET** is also among top 5 contributors differentiating ASD from NT.

### 2- Generating ASD Subtypes and Validating them

The classifier that generated three subtypes includes 5 domains of NEO-PI-r, and RMET test scores. We used several clinical, behavioral and neuroimaging measures that are not included in the initial model to validate the putative subtypes. We considered the model successful if the subtypes differed in at least one of the clinical diagnostic measures or on one of its sub measures.

#### 2.1. Behavioral differences between subtypes on measures that are part of the classifier

**A- NEO-PI-r personality test:**

We found significant differences between groups in **Neuroticism** domain (F (3, 102) = 16.508, P < 0.001, eta2 = 0.327) and its 6 facets (Fig. 2A, 2E) with Subtypes 1 and 3 having higher scores and higher scores in *Anxiety*, *Depression*, *Self-Consciousness*, *Vulnerability* compared to NT and Subtype 2 (P < 0.001; P < 0.005). Subtype 3 shows higher “*Angry/Hostility*” compared to Subtype 2 (P =0.041) and to NT (P < 0.001) and displays higher *Impulsiveness* compared to Subtype 2 (P =0.012) but not compared to NT (P = 0.207).

**Figure 2:**
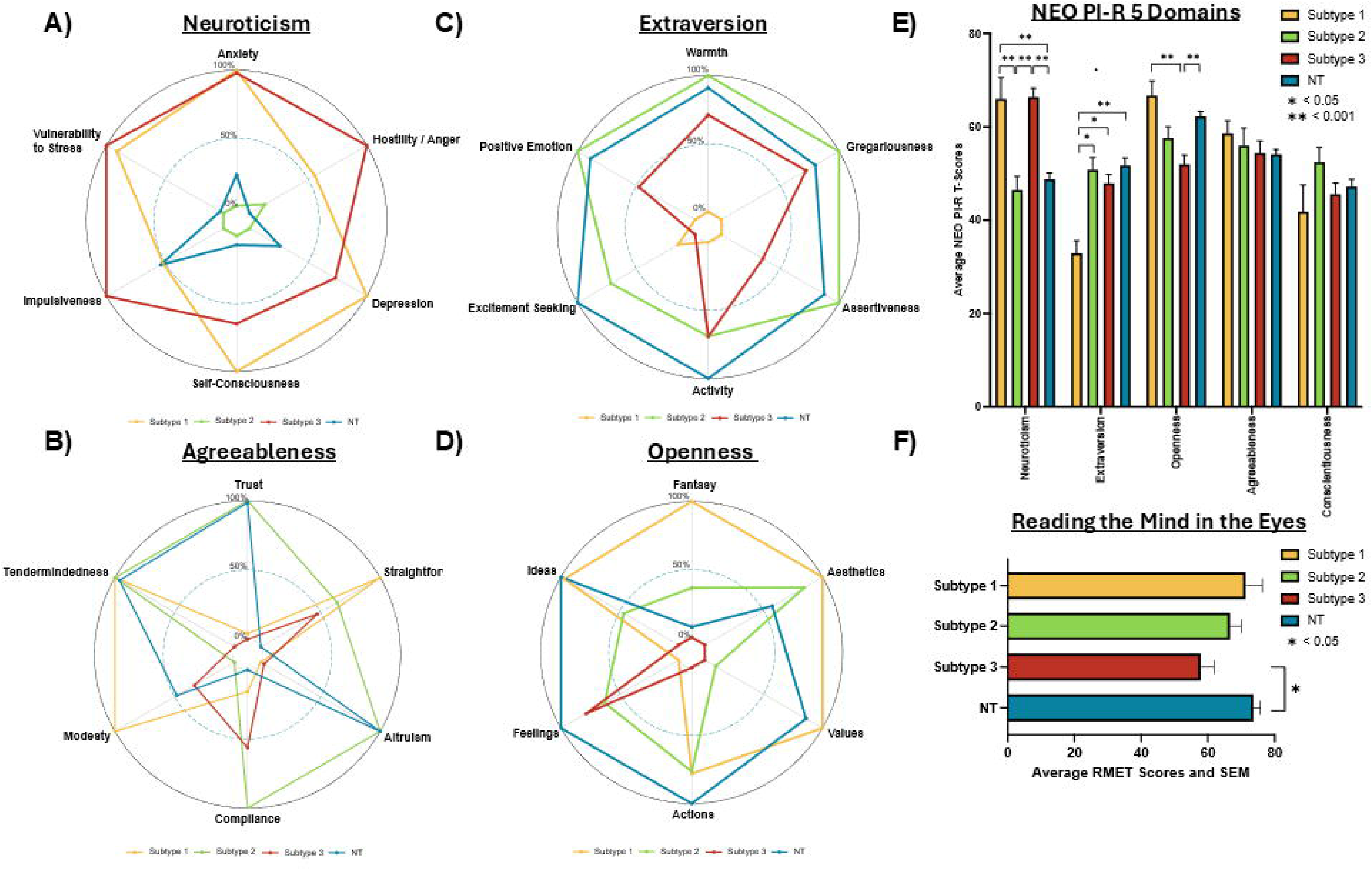
Model inputs - NEO-PI-r Facets and RMET Scores. Figure 2A-D descriptively illustrates the NEO-PI-r Facets that were significant compared between the subtypes and NT; A) is for Neuroticism, B) Agreeableness, C) Extraversion, and D) Openness (note: the Conscientious radar plot is in the supplemental section). Figure 2E is the bar graph comparing the main NEO-PI-r domain scores between the subtypes and NT. Lastly, 2F compares the RMET test between the subtypes and NT. Error bars figure E and F are computed based on the standard error of the mean for each group. * P< 0.05; ** P< 0.001.

There is a significant difference between groups in **Extraversion** (F (3, 102) = 7.624, P < 0.001, <u>eta2 = 0.183</u>), and its 6 facets (Fig. 2C, 2E) with <u>Subtype 1 having lower levels of Extraversion and Warmth compared to Subtype 2 (P = 0.006), Subtype 3 (P = 0.012) and to NT (p < 0.001).</u> Subtype 1 participants displayed lower *Gregariousness, Assertiveness, and Positive Emotions* compared to Subtype 2 and NT (P < 0.05), and lower *Activity Level* compared to NT (P = 0.002). Subtype 3 had lower *Excitement Seeking scores* compared to NT (P = 0.04).

Differences between groups were also noted for the domain of ***<u>Openness</u>*** *(F* (3, 102) = 7.632, P < 0.001, eta2 = 0.183), and *differences in facets of Actions (F* (3, 101) = 6.655, P < 0.001, eta2 = 0.165)*, Ideas (F* (3, 101) = 6.371, P < 0.001, eta2 = 0.159)*, and Values (F* (3, 102) = 16.824, P < 0.001, eta2 = 0.331) (Fig. 2D, 2E). <u>Subtype 3 shows lower scores in Openness, *Values*, and *Ideas*</u> compared to Subtype 1 and NT (P < 0.05), and in *Action* compared to NT (P < 0.001). Subtype 2 also had lower scores in *Values* compared to Subtype 1 and NT (P = 0.001).

For *Straightforwardness* facet, there are significant differences between subtypes and NT (F (3, 102) = 5.168, P = 0.02, eta2 = 0.132) with <u>Subtype 1 having significantly higher scores than NT</u> (P = 0.007) (Fig. S1, 2E).

**B- The *Reading the Mind in the Eyes*** test (RMET) differed significantly among groups, with Subtype 3 showing significantly lower RMET scores compared to NT (F (3, 66) = 3.914, P = 0.012, eta2 = 0.151; P = 0.009). No other differences were found following Bonferroni corrections (Fig. 2F).

**C- <u>SCL-90-R</u>**: There are significant differences between groups on all sub scores (Fig. S2, 3C) see Supplementary Result section for detailed statistics), with Subtypes 1 and 3 showing higher scores on *Obsessive-Compulsive, Interpersonal Sensitivity, Depression, Anxiety, Phobic Anxiety, Psychoticism, Global Severity Index,* and *Positive Symptom Total* compared to Subtype 2 and NT (P <0.05). Subtype 3 shows higher Hostility, Paranoid Ideation, Positive Symptom Distress, and Somatization compared to Subtype 2 and NT (P < 0.05). Subtype 1 shows higher Paranoid ideation and Positive Symptom Distress Index only compared to NT (P < 0.05).

**D- <u>The *Autism Quotient*</u>** was significantly higher for Subtype 1 compared to Subtype 2 and NT (F (3, 100) = 26.352, P < 0.001, eta2 =0.442; P < 0.05) but not compared to Subtype 3 (P = 0.281). Subtypes 2 and 3 have higher AQ scores in comparison to NT (P < 0.05) (Fig. 3A).

**Figure 3:**
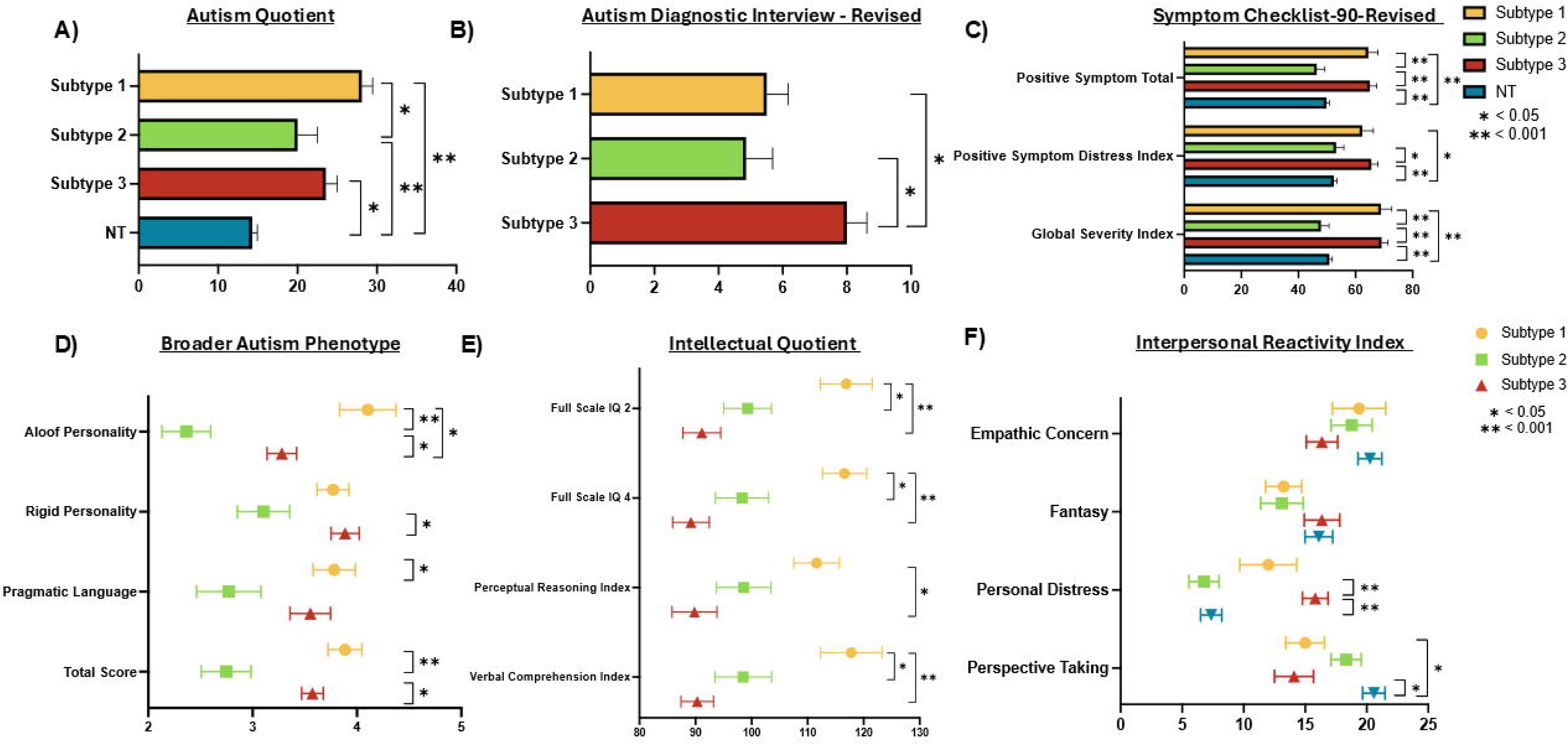
Behavioral Validation Features. Figure 3A is the Autism Quotient compared to NT and Controls. 3B is the ADI-R for the Repetitive Behaviors Score compared between subtypes. 3C is the SCL-90-R is the abbreviated SCL figure which included Positive Symptom Total, Positive Symptom Distress index, and Global Severity Index Compared between Subtypes and NT (Note the full version of the SLC-90-R graph can be found in the supplemental section). Figure 3D is the Boarder Autism Phenotype Box Plots compared between subtypes. Figure 3E is the Intellectual Quotient between subtypes. Figure 2F is the Interpersonal Reactivity Index compared across subtypes and NT. Figure 2A-C error bars are computed based on the standard error of the mean for each group. Figure 2D-F standard error bras are computed based on the standard error of the mean for each group. * P< 0.05; ** P< 0.001.

**Figure 4:**
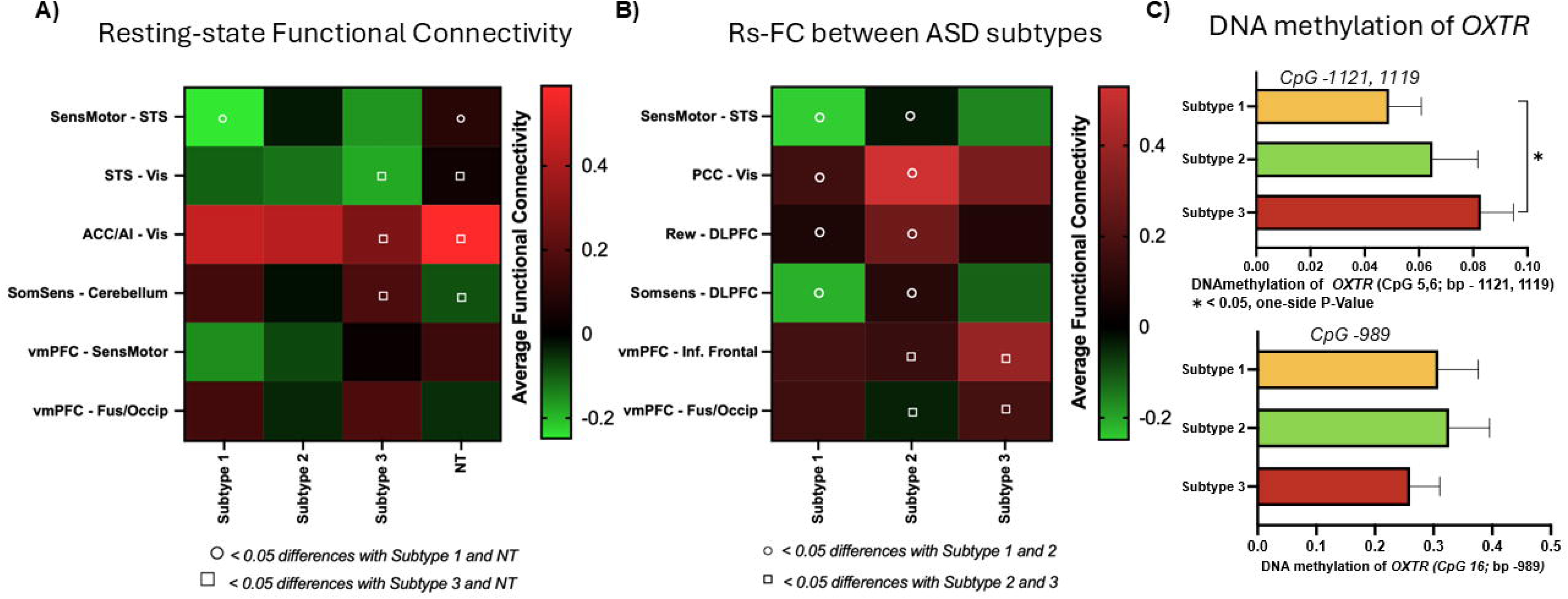
Biological Validation Features. Figure 4A is a heatmap comparing the resting state functional connectivity between subtypes and NT. The “circle” indicates statistically significant difference between subtype 1 and NT; the “square” indicates statistically significant difference between subtype 3 and NT. B is a heatmap comparing the resting state connectivity between subtypes only. The “circle” indicates statistically significant difference between subtype 1 and 2; the “square” indicates statistically significant difference between subtype 1 and 3. The colors of the cells are shaded according to average functional connectivity levels, with *red* indicating higher connectivity and *green* lower connectivity. Figure C is the CpG methylation for Chromosome 5,6 (b.p. – 1121, 1119). Figure D is the CpG methylation for Chromosome 16 (b.p. – 989). The error bars in figure C and D were derived from the standard error computed from the mean. * P< 0.05.

**E- <u>The *intelligence quotient*</u>** is statistically different between groups on *all scores with* Subtype 1 showing significantly higher scores on the *full-scale IQ-4 composite*, on the *Full-Scale IQ-2 composite*, and on the *Verbal Composite Index* in comparison to Subtypes 2 and 3 (P < 0.05). For the *Perceptual Reasoning Composite index,* Subtype 1 shows higher scores compared to Subtype 3 (P < 0.05) (Fig. 3E).

**F- <u>The *BAPQ*</u>** has significant differences in BAPQ total scores (F(2, 24) = 11.536, P < 0.001, eta2 = 0.490) and in *Aloofness, Rigidity, Pragmatic Language*. Subtypes 1 and 3 show higher average total BAPQ scores compared to Subtype 2 (P < 0.005). Subtype 1 showed higher *Aloofness* in comparison to both Subtypes 2 and 3 (P < 0.05) and higher *Pragmatic Language deficits* compared to Subtype 2 (P < 0.05). Subtype 3 shows higher *Rigidity* scores on BAPQ compared to Subtype 2 (P < 0.05) (Fig. 3D).

**G- <u>The IRI</u>** showed significant differences in personal distress and perspective taking between 3 subtypes and NT (F (3, 43) = 11.541, P < 0.001, eta2 = 0.446; F (3, 43) = 6.453, P = 0.001, eta2 = 0.310), with Subtype 3 showing higher personal distress scores compared to Subtype 2 and NT (P < 0.001). Perspective taking for Subtypes 1 and 3 is lower than NT (P < 0.05) (Fig. 3F).

### 2.2. Clinical validation

ADI-R yielded significant differences between subtypes for the ADI-R repetitive behaviors and restricted interests (F (2,30) = 5.941, P = 0.007, eta2 = 0.284). No differences were found for the other sub scores (social, communication) or total score (P > 0.05). Subtype 3 has significantly higher ADI-R repetitive behaviors than Subtypes 1 and 2 (P < 0.05) (Fig. S4). No significant differences were found for the ADOS-2 comparison scores.

### 2.3. Biological validation of the 3 putative subtypes

#### A- Resting-state Functional Connectivity results

For the analysis that included 3 ASD subtypes and NT, we found significant differences among groups in the rs-Functional Connectivity between Sensorimotor network and theory of mind network (STS) (F (3, 41) = 4.289, P = 0.010, eta2 = 0.239); between Sensorimotor network and Ventromedial PFC (or default network) (F (3,41) = 3.086, P = 0.038, eta2 = 0.184); between Theory of Mind (STS) and Visual network (F (3, 41) = 3.582, P =0.022, eta2 = 0.208); between Ventromedial PFC (or default network) and Fusiform Occipital network (F(3, 41) = 2.969, P = 0.043, eta2 = 0.178); between Empathy/salience network (ACC, AI) and Visual cortex (F (3, 40) = 3.442, P = 0.026, eta2 = 0.205); and between Somatosensory network and Brainstem/Cerebellum network (F (3,41) = 4.4104, P = 0.012, eta2 = 0.231); (See Fig 5A, 5B). <u>Subtype 1 showed lower rsFC between Sensorimotor and STS compared to NT (P = 0.021). Subtype 3 exhibited *hypoconnectivity* between STS and visual cortex, between ACC, AI and visual cortex and has *hyperconnectivity* between somatosensory and Brainstem/Cerebellum network compared to NT (P<0.05).</u>

For the analysis that includes only 3 subtypes of ASD without NT, Subtype 1 showed lower rsFC compared to Subtype 2 between Sensorimotor network and STS (t (14) = -2.375, P = 0.032, Cohen’s d = -1.197); between PCC and visual cortex (t (14) = -3.720, P = 0.002; Cohen’s d = - 1.875); between Reward network to dlPFC areas (t (14) = -2.296, P = 0.038, Cohen’s d = -1.157); and between Somatosensory and dlPFC ( t(14) = -2.558, P = 0.023, Cohen’s d = -1.289).

Subtype 3 showed hyperconnectivity between vmPFC (default mode network) and IFG compared to Subtype 1 (t (16) = -2.266, P = 0.038, d = -1.096) and Subtype 2 (t (18) = -2.608, P = 0.009, d = -1.172); and a hyperconnectivity between vmPFC and fusiform/occipital areas compared to subtype 2 (t(19) = -2.232, P = 0.038, d =-0.984).

#### B- DNA methylation of the oxytocin receptor gene

Subtype 3 had higher *OXTR* DNA methylation at CpG 5.6 site compared to Subtype 1 (t (22) = - 1.953, P = 0.032 with one-sided p value, Cohen’s D = -0.809). No differences were found between Subtypes 1 and 2 (t(16) = -0.802, P = 0.217, Cohen’s D = -0.381) nor between Subtypes 2 and 3 (t (20) = -0.885, P = 0.193, Cohen’s D = -0.392). No differences were found for CpG 16 site between subtypes (Fig.5C, 5D), however, the ANOVA between subtypes and NT was significant (F(3, 87) = 5.092, P = 0.003, eta2 = 0.265) in line with what was found previously (Andari et al., 2020). Subtype 2 was shown to have significantly more methylation than NT (P = 0.003).

### 2.4 Personalized drug treatments – Intranasal oxytocin (Syntocinon spray Novartis) as one potential drug treatment

We investigated interactions among the different factors (subtypes of ASD, all doses of IN-OXT, and rsFC networks) (see Table 1) and found that Subtype 1 seems to be substantially affected by IN-OXT, as the high dose of 48IU increased significantly rsFC <u>between PCC and Cerebellum</u> <u>compared to placebo</u> (P = 0.009); and 48IU of IN-OXT decreased significantly rsFC between <u>Cerebellum/Brainstem and Parietal lobe</u> compared to 24 IU (M = 0.077, SE = 0.071) and placebo (M = 0.095, SE = 0.071) (P =0.038, P = 0.021). There is a significant interaction between subtypes and dosage for the foregoing FNs (F (6, 74.107) = 3.850, P = 0.002); F (6, 75.039) = 2.437, P = 0.03) (see Supplementary Material for further results comparing high dose of IN-OXT to placebo).

| Brain Region Network | DF Within | DF Between | F-Stat | P-Value | Subtype | Dosage 1 | M | SE | Dosage 2 | M | SE | Direction | Pvalue |
| --- | --- | --- | --- | --- | --- | --- | --- | --- | --- | --- | --- | --- | --- |
| Parietal and Frontal Lobes to Visual Lobe | 2 | 25 | 3.747 | 0.038 | 1 | 48 IU | -0.249 | 0.091 | Placebo | -0.033 | 0.091 | Decrease | 0.026 |
| - | - | - | - | - | 2 | 48 IU | -0.144 | 0.079 | Placebo | 0.06 | 0.079 | Decrease | 0.018 |
| Parietal and Frontal to Anterior Cingulate Cortex (Pain) | 2 | 25 | 4.203 | 0.027 | 1 | 48 IU | -0.155 | 0.111 | Placebo | 0.104 | 0.111 | Decrease | 0.021 |
| Cerebellum to Posterior Cingulate Cortex | 2 | 25 | 6.648 | 0.005 | 1 | 48 IU | 0.406 | 0.1 | Placebo | 0.1 | 0.074 | Increase | 0.001 |
| - | - | - | - | - | 3 | 48 IU | 0.423 | 0.056 | Placebo | 0.294 | 0.056 | Increase | 0.049 |
| Cerebellum Brainstem to Parietal and Frontal Lobes | 2 | 49 | 4.789 | 0.013 | 2 | 48 IU | 0.132 | 0.042 | Placebo | -0.047 | 0.042 | Increase | 0.004 |
| Cerebellum Brainstem to Sensorimotor | 2 | 49 | 5.071 | 0.01 | 1 | 48 IU | -0.096 | 0.067 | Placebo | 0.137 | 0.067 | Increase | 0.016 |
| Cerebellum Brainstem to Parietal Lobe | 2 | 25 | 4.992 | 0.015 | 1 | 48 IU | -0.173 | 0.072 | Placebo | 0.095 | 0.072 | Decrease | 0.0009 |
| Superior Temporal Sulcus to Visual Lobe | 2 | 25.002 | 4.545 | 0.021 | 3 | 48 IU | 0.14 | 0.082 | Placebo | -0.055 | 0.082 | Increase | 0.002 |

## Discussion

In this study, we first predicted ASD diagnosis using a comprehensive personality test that encompasses domains of sociability and negative valence and using a theory of mind test that measures empathic capacities. We successfully created a random forest model that accurately classifies ASD and NT with an AUC of 0.82 ± 0.14. The SHAP feature contribution analysis indicated that Values, Vulnerability, Extraversion, Activity, Straightforwardness, Neuroticism, Excitement Seeking, and RMET are among the most important diagnostic predictors.

Second, we were also able to generate subtypes of ASD using the NEO-PI-r and RMET task, and we further validated these subtypes using clinical and biological measures. While Subtype 2 did not show distinct features, Subtypes 1 and 3 differed significantly on several features from Neurotypicals.

**Subtype 1 or** “***Negative Affect with Social Aloofness or NASA Subtype***,” displayed <u>high Neuroticism</u>, high Anxiety, Depression, and Vulnerability, high scores on Symptom Checklist and negative affect, as well as <u>low scores on Extraversion</u>, lower warmth, high Aloofness based on BAPQ results, and more deficits in pragmatic language, signaling significant increase in negative valence as well as lower sociability. The NASA subgroup has significantly higher IQ scores than the other groups and displays more straightforwardness meaning it can be considered as a more direct subgroup with blunt and honest ways of communicating.

In terms of biological underpinnings, we found that NASA participants showed lower rsFC between sensorimotor network and Superior Temporal Sulcus (STS) network and hypoconnectivity between STS and visual cortex and between ACC/Insula network and visual cortex. These networks are heavily involved in cognitive and emotional empathy as well as salience processing [59–67].

The FC between STS and visual cortex has been documented to be involved in eye contact and face processing and therefore the NASA subgroup might be more deficient in eye gaze and processing of social cues such as facial expressions [68]. This corroborates the behavioral findings with higher AQ scores and higher aloofness scores in this subgroup. Based on rsFC data, the NASA subgroup likely has deficits in sensorimotor processing. A reduction in functional connectivity between ACC/AI and visual cortex has been reported to be directly linked to sensory problems in ASD [69]. Hypoconnectivity between STS and sensorimotor networks and between somatosensory and dlPFC are all indicators of a disruption in multisensory integration of information. We do not have behavioral results regarding sensory problems in this study to back up the brain functional data, but future studies should include sensory evaluations to better characterize NASA subgroup of ASD. In addition, the NASA subgroup showed hyperconnectivity between somatosensory network and brainstem/Cerebellum which is also related to prediction of sensory consequences of self-produced touch, and which could be related to a reduced sensitivity [70]. There is growing evidence in the literature discussing hyperconnectivity between subcortical and cortical areas in ASD [21,71,72].

The NASA group also showed hypoconnectivity between default mode network (PCC in particular) and visual cortex which is related to self-reflection, introspection, self-awareness, and is directly linked to social deficits that this subtype struggles with [73–75]. A reduction in rsFC between reward network that includes nucleus accumbens, and dLPFC has been previously linked to anhedonia and this is in line with the lack of warmth and emotional detachment or social avoidance that this group displays [76–78].

**Subtype 3 or “Neurocognitive and Affect Dysfunctions with Resistance to change or NADR Subtype”** <u>showed higher levels of Neuroticism</u> and significantly <u>lower Openness</u> and lower scores on Values and Ideas indicating that these individuals are less open to experience and prefer familiarity with routines over novelty. They have higher scores on Anxiety, Depression, but also on Anger and Impulsivity, along with higher personal distress on IRI which signal emotion dysregulation. This subtype has higher scores on rigidity in the BAPQ test, and higher scores on ADI-RRB, indicating more resistance to change and the presence of restricted interests.

In addition to higher reports of repetitive behaviors and restricted interests when young (based on ADI-R), this subtype displays higher connectivity between default mode (vmPFC) and inferior frontal cortex and occipital areas. This is in line with previous reports describing hyperconnectivity in ASD that can be linked to repetitive behaviors and restricted interests [79–84]. The default mode network also included some reward areas. Hyperconnectivity between default mode network and other networks have been linked to social anxiety disorder [71,85]. This subtype also shows hypermethylation in the *OTXR* gene on a CpG site that was previously linked to autism deficits and lower scores on the RMET task which can be related to deficits in cognitive empathy. This subtype also displays lower IQ scores which could signify more cognitive difficulties alongside higher rigidity and negative affect.

In terms of differential reactivity to oxytocin intake, when analyzed in a mixed model with different doses of OXT, the **NASA** subtype seems to be the most affected by IN-OXT in which the highest dose increased connectivity between PCC and Cerebellum but decreased connectivity between the Brainstem and Cerebellum network (that was found hyperconnected in NASA) and Parietal Network. Connectivity between PCC and Cerebellum is related to cognitive and affective and self-referential processes and IN-OXT has been previously shown to enhance this network in women with supportive maternal care [74,86–88]. Hyperconnectivity between Brainstem/Cerebellum and the Parietal cortex has been associated with depression and anxiety increase (Gasper et al., 2025) and therefore the fact there was a dose-dependent decrease in NASA with IN-OXT, it could signify that this drug can have promising action on negative valence in addition to social cognition in NASA [89–92]. It is therefore possible that patients fitting the NASA profile would benefit from OXT treatment or similar treatment sharing similar mechanisms of action given that these individuals were characterized by lower scores in sociability, warmth and gregariousness and lower functional connectivity between networks involved in social salience and empathy, such as ACC-Insula, and Superior Temporal Cortex. It is possible that Subtype 3 or **NADR, characterized** by higher levels of DNA methylation of the *OXTR* gene, are less susceptible to change with this treatment but could benefit from other treatments. Also, **NADR** is characterized by higher levels of rigidity and lower Openness, which was previously linked to more resistance to treatment.

A recent study of Zhao, et. al., 2024 attempted to check if IN-OXT can moderate one subtype of ASD more effectively than another and showed that a subtype of ASD with the highest levels of autism severity showed a decreased in ADOS scores and greater interest in eye-region of emotional faces following IN-OXT [93].

Limitations of the study include small sample size, as well as its focus on males. Replication studies are needed in women and men. We did not have behavioral data for the OXT trial that can support fMRI rs-FC data and therefore future work needs to include targeted behavioral measures for OXT treatment. Inclusion of sensorimotor measures will be essential to better characterize subtypes. Our data did not include neuroimaging rsFC for Neurotypicals and therefore we were not able to run the classifier with behavioral and neuroimaging data. Future models can include functional brain data in the classifier to adopt a multimodal approach in predicting ASD diagnosis and in generating ASD subtypes.

## Data Availability

All data produced in the present study are available upon reasonable request to the authors

## Author contributions

C.D.V. analyzed the data and wrote the manuscript. J.H-K. analyzed and co-wrote the manuscript. K.G. analyzed the data and revised the manuscript. M.M. collected data and revised manuscript. O.O collected data and revised it. J.F.C. collected data and co-wrote the manuscript. L.Y. was the PI on the grant that helped collect the original data at Emory. He is currently deceased and could not edit the manuscript. E.A. conceived and designed the study, collected data, analyzed the data, interpreted the data, and wrote the manuscript.

## Funding and Acknowledgment

The work is also supported by ProMedica Health System Foundation grant to EA (Autism and Social Neuroscience, index number 207007). The collection of data was supported by NIH grants P50MH100023 to LJY, P51OD11132 to Emory National Primate Research Center. The project was also supported by National Institutes of Health/National Center for Advancing Translational Sciences grant UL1 TR002378 and the Georgia Clinical and Translational Science Alliance. We thank all investigators who helped collect data from the main project at Emory University including Dr. James Rilling and Dr. Alica Smith. We would also like to thank Gwen Hoffman for proofreading this manuscript and Dr. Ali Imami for their assistance. Finally, we would like to thank Dr. Daniella Gamboa Pabon for her support.

