## Supplemental file for "Generating Biologically Relevant Subtypes of Autism Spectrum Disorder with differential responses to Acute Oxytocin Administration in a Randomized Trial using Random Forest Models and K-means Clustering"

Supplementary Methods

### Methods and Materials

#### Subjects

Subjects were recruited from Emory University as part of the Autism Oxytocin Brain Project. All subjects were adult males between 18 and 45 years old with an Intelligence Quotient (IQ) greater than 70. We initially recruited 74 neurotypical subjects and 50 with ASD to conduct a series of behavioral tasks and saliva collection for DNA methylation analysis of the oxytocin receptor gene as part of the Emory IRB#63466. 10 ASD subjects were excluded because of eligibility criteria which led to a N of 40 for ASD sample size for behavioral data collection. One subject withdrew from the study and therefore the total number of ASD was 39 for the actual analysis. Three NT individuals did not complete the NEO-PI-r which led to 71 as the total number of NT for data analysis.

Only 30 adults with ASD completed the clinical trial with oxytocin and fMRI (resting state as one of the tasks) as part of the Emory IRB#9455. We also included a new set of NT (17 male controls for IQ, age, and race) for the fMRI results (see Supplementary file and previous publication) [1]. 2 ASD subjects did not have data for the NEO-PI-r personality data and therefore were excluded from the analysis. This gives us a sample of 28 total ASD for the analysis.

Finally, the *Autism Diagnostic Observation Schedule* (ADOS) and *Autism Diagnostic Interview* tests were used to confirm ASD diagnosis. These tests were used to validate the ASD subtypes and to detect any measurable differences between them [2,3].

#### Behavioral tests, surveys, and clinical assessments

All subjects were tested with the personality test NEO-PI-r[4], Reading the Mind in the Eyes Test (RMET) [5,6], Symptom Checklist 90 (SCL-90-R) [7,8], Intelligence Quotient (IQ) [9,10], Social Responsiveness Scale (SRS-2) [11,12], Broader Autism Phenotype Questionnaire (BAPQ) [13], and Autism-Spectrum Quotient (AQ) [14]. Those who were selected for the clinical trial also completed the Interpersonal Reactivity Index (IRI) as a measure of empathy including perspective taking, fantasy, empathic concern, and personal distress [15].

**Machine Learning and Statistical Methods**

*Classifying ASD and NT*

RMET and NEO-PI-r tests were compiled for both Autism and Neurotypical subjects. All machine learning and statistical analysis was conducted in Python [16]. Missing data points were imputed using the missForest from the missingpy package [17]. A hyper parameterized gridsearch using SciKit-Learn was then conducted to find the most optimal parameters for a Random Forest Tree Model comparing the ASD and NT Accuracy, Specificity, ROC, and AUC were calculated for the Random Forest Tree model [18]. Shapley Additive exPlanations values (SHAP) were calculated using the SHAP package from the Random Forest Tree Model to calculate the relative contributions of each feature and most relevant features [19,20].

**Methods and Materials**

**Subjects**

Subjects were recruited between 2012 to 2019 at Emory University as part of the Autism Oxytocin Brain Project. All subjects were adult males and between 18 and 45 years old with an Intelligent Quotient greater than 70. We initially recruited 74 neurotypical individuals and 50 subjects with

ASD to conduct a series of behavioral tasks and saliva collection for DNA methylation analysis of the oxytocin receptor gene as part of the Emory IRB#63466. 10 of ASD subjects were excluded because of eligibility criteria which leads to a N of 40 for ASD sample size for behavioral data collection. Around 70% of ASD participants were Caucasian, 20% were African American, and 7.5% more than one race, and 2.5% were unknown. One subject withdrew from the study and therefore the total number of ASD was 39 for the actual analysis. Three NT individuals did not complete the NEO-Pi-r which led to 71 as total number of NT for data analysis.

Of those, only 30 adults with ASD continued the clinical trial with oxytocin and fMRI (resting state as one of the tasks) as part of the Emory IRB#9455. ASD were recruited from the Emory Autism Center with Dr. Cubells (J.C., psychiatrist on the study) confirming ASD diagnosis clinically and with Dr. Opal Ousley and Dr. Michael Morrier confirming ASD diagnosis with ADOS-2 and ADI-R which are gold standard measures. Therefore, we only included 30 subjects for rsFC for the validation analysis and for the drug target engagement section. We also included a new set of NT (17 male controls for IQ and for age and race) for the fMRI results as NT from the first study did not continue with the clinical trial (Please see details for experimental procedure for the clinical trial in Andari et al., 2025) [1].

2 ASD subjects did not have data for the NEO-PI-r personality data and therefore were excluded from the analysis. This gives us a sample of 28 total ASD for the analysis.

Data used here in this study is also used in part in Andari et al. 2025 (NPP) but for different purposes [1]. In the original study, we showed how oxytocin administration affects the brain rsFC using machine learning approaches. In this study, we are using additional data to generate subtypes of ASD and then use neuroimaging rsFC data on the different subgroups to see if they are significantly different.

**Behavioral tests, surveys, and clinical assessments**

All subjects were tested with the NEO-PI-R, RMET, SCL-90-R, IQ, SRS, BAPQ, and AQ. Those with autism were also tested with ADOS and ADI-R to confirm their diagnosis.

The *NEO-PI-R* test was used to measure relative personality across domains including 'Openness' (active imagination, aesthetic sensitivity, and intellectual curiosity), 'Conscientiousness' (tendency to show self-discipline, act dutifully, and aim for achievement), 'Extraversion' (tendency to enjoy human interactions, enjoy time spent with people, and find less reward in time spent alone'), 'Agreeableness' (tendency to be compassionate and cooperative), and 'Neuroticism' (tendency to experience negative emotions, emotional instability) [4].

The *Reading Mind in the Eye Test* was used to measure the ability to identify emotions from images of facial expressions. RMET is intended to test theory of mind abilities [5,6].

The *Symptom Checklist-90 Revisited* or SCL-90-R is a screening test used to self-report psychological symptoms associated with somatization, obsessive-compulsive traits, interpersonal sensitivity, depression, anxiety, and hostility, phobic anxiety, paranoid ideation, and psychoticism psychological conditions in addition to autism or lack thereof. This data was compared to the neurotypicals to see if there was a significant difference [7,8].

The *Intelligence Quotient* (IQ) was used to test subjects across different cognition domains and compare them to the prescribed subtypes and neurotypicals. This measu.re allows us to measure relative intelligent levels and performance on different tasks [9,10].

The Interpersonal Reactivity Index (IRI) was used to measure perspective taking, fantasy, empathic concern, and personal distress [15].

The *Social Responsiveness Scale* (SRS) was used to measure social communication ability. This scale allows us to test the relative variability between socialization levels and allows us to draw conclusions between the different phenotypes [11,12].

*Autism Quotient* (AQ) test is a self-administrated test that is to score autism features based on five key features: Social Skills, Attention Switching, Attention Deficit, Communication, and Imagination [14].

The *Broad Autism Phenotype Questionnaire* (BAPQ) uses a set of personality and language characteristics to measure the autism phenotype across the categories of social deficits, stereotyped repetitive behaviors, and social language [13].

Both NT and ASD were measured with these tests. The ASD subtypes scores were compared to NT to ensure the validity of the subtypes. They were also compared against each other to see if there were differences between the subtypes [14,21].

Finally, the *Autism Diagnostic Observation Schedule* (ADOS) and *Autism Diagnostic Interview* tests were used to confirm ASD diagnosis. These tests were used to validate the ASD subtypes and to detect any measurable differences between them [2,3].

**Methylation Methods**

Saliva samples were collected using Salivette® (SARSTEDT) and kept chilled on ice for up to 2 h before being stored at −80 °C until the day of separation. DNA was extracted from the Salivette® swab using the QIAamp mini kit (Qiagen, Hilden, Germany) and was quantified with PicoGreen® (Quant-iTTM PicoGreen® dsDNA Assay Kit, Thermo Fisher Scientific Inc., Waltham, MA). The average yield from 100 samples was 2.7 µg/swab. One microgram of DNA was treated with sodium bisulfite using the EpiTect Fast Bisulfite Kit (Qiagen, Hilden, Germany).

The *OXTR* gene (chr3: 8792095 to 8811300; GRCh37/hg19 build) was interrogated using EpiTYPER (MassARRAY system; Agena Bioscience, San Diego, CA) according to the manufacturer's instructions. We aimed to target a specific 406-bp region (chr3: 8810719–8811124) termed MT2 [22], located in a CpG island, as it is the regulatory region related to transcription of the gene. We selected the EpiTYPER platform to cover almost all CpG sites (21/27 CpGs in MT2) (see Supplementary material).

For each participant, the *OXTR* methylation ratios were retained and used as the criterion variable in subsequent statistical analyses. The reproducibility and sensitivity of the *OXTR* EpiTYPER assay product were assessed using commercially available standards (EpiTect control DNA, Qiagen) run in triplicate and measured as previously shown [23].

**Resting State fMRI acquisition and pre-processing**

30 of those with ASD and 18 of the NT, underwent a resting-state fMRI.

Each patient took part in an 8-minutes rsfMRI scan followed by other tasks that will be reported in future publications. Patients were instructed to keep their eyes open and look at a fixation cross that is presented on a screen in front of them during the resting-state scans. MR images were acquired on a 3T Siemens Magnetom Trio TIM scanner with a 32-channel receiver array head coil. Blood oxygenation level dependent (BOLD) contrast resting state fMRI (rsfMRI) scans were acquired using a simultaneous multi-slice (SMS) slice-accelerated gradient echo EPI sequence (Feinberg et al. 2010; Moeller et al., 2010) with imaging parameters: field-of-view (FOV) = 220mm; repetition time (TR)/echo time (TE) = 1000ms/26ms; flip angle (FA) = 60˚; 74 X 74 matrix size; 72 axial slices of 2mm width covering the whole brain; multiband acceleration factor = 6; in-plane GRAPPA acceleration factor = 2; 280 scan volumes. Two five-volume spin-echo conventional EPI sequences with same image resolution and coverage and TE as the GE EPI sequence described above were also acquired, one with the same direction of phase-encoding (AP), and one with reversed PE direction (PA), were also acquired to assist with distortion correction of GE BOLD EPI data. A whole-brain 3D T1-weighted MPRAGE sequence (FOV = 230 mm; TR/TI/TE/FA = 2250 ms/900 ms/3 ms/9°; 0.9 mm x 0.9 mm x 1 mm resolution) provided anatomic detail. Foam padding was provided to minimize subject-motion during the scans.

Data analysis was performed using the AFNI and FSL [24] software packages, the GIFT toolbox [25] for GICA, and scripted programs written in Matlab.

The rsfMRI voxel time-series were corrected for geometric distortions, temporally shifted to account for differences in slice acquisition times, 3D volume registered to a base volume to account for global rigid motion and spatially smoothed with an isotropic Gaussian filter (FWHM = 6 mm). The resultant rsfMRI time-series were co-registered to the T1-weighted high-resolution anatomic scan using the well-established affine boundary-based registration algorithm [26] and spatially normalized to the MNI152 template with the warp computed from alignment of the high-resolution 3D anatomic to the MNI152 template with a nonlinear registration algorithm.

**Oxytocin Clinical Trial**

The study consisted of a randomized, placebo-controlled, double-blind, between-subjects design to reduce the learning effect of repetitive sessions. Participants received a single intranasal dose of either 48 IU, 24 IU, 8 IU of oxytocin (Syntocinon Spray; Novartis; three puffs per nostril) or placebo 30 min before the start of experiments in the MRI scanner. We pre-calculated that an additional interval of 10–15 min would be necessary for installing the subject inside the scanner and starting the paradigm, which would add up to 40–45 min, the necessary duration for Syntocinon spray effects on the brain. Inside the scanner, Patients were instructed to keep their eyes open and look at a fixation cross that is presented on a screen in front of them during the resting-state scans.

#### Oxytocin Clinical Trial MRI acquisition

MR images were acquired on a 3T Siemens Prisma-Fit scanner with a 64-channel receiver array head + neck coil. BOLD contrast rsfMRI scans were acquired using a conventional EPI sequence with FOV = 192 mm, TR/TE/FA = 3000 ms/25 ms/90°; fifty-eight 2.4-mm-thick oblique slices; 1.5 mm × 1.5 mm in-plane resolution. There were 160 measurements in each rsfMRI scan.

#### Oxytocin Clinical Trial Data analysis

The rsfMRI data were analyzed with standard preprocessing pipeline including image distortion correction, slice-time correction, 3D volume registration, and spatial normalization to MNI152 template. The preprocessing pipeline (see Supplementary material for more details) included an added motion artifact reduction step conducted through the independent component analysis (ICA)-AROMA technique [27] ICA was performed on temporally concatenated data of the whole subject cohort using the GIFT software [28]. Whole-brain group ICA spatial maps (SMs; with component strength expressed as *t*-scores), as well as corresponding IC time-courses (TCs) for each subject were obtained through back-projection [28]. The functional classification of the ICs were performed with image based meta-analyses conducted with the publicly available Neurosynth meta-analyses database [29]; utilizing the well-established correspondence between resting-state networks and task-related brain networks [30]. Functional network connectivity (FNC) was assessed between pairs of IC networks that can be unambiguously classified as representing specific brain functions or functional domains, through cross-correlation coefficient (CC) of their respective TCs.

### Machine Learning and Statistical Methods

All Analysis Scripts are available publicly on Github. SPSS output is also available on github as a PDF.

The machine learning and initial statistical analysis was conducted in Python 3 [31].

*Classifying ASD and NT*

RMET and NEO-PI-R tests were compiled for both Autism and Neurotypical subjects. All machine learning and statistical analysis were conducted in Python [16]. Missing data points were imputed using the missForest from the missingpy package [17]. A hyper parameterized gridsearch using SciKit-Learn was then conducted to find the most optimal parameters for a Random Forest Tree Model comparing the ASD and NT Accuracy, Specificity, ROC, and AUC were calculated for the Random Forest Tree model [18]. Shapley Additive exPlanations values (SHAP) were calculated using the SHAP package from the Random Forest Tree Model to calculate the relative contributions of each feature and most relevant features [19,20].

*Clustering ASD*

SHAP values of the individual subjects were subsequently calculated. The log odds of the ASD and NT were then calculated for each SHAP frame and then normalized [19,20]. NT were then dropped from subsequent analysis. The ASD Normalized SHAP and Log Odds were visualized together status as Principal Component Analysis and clustered together using K-Means Clustering from the Sci-Kit Learn Package. The optimal number of clusters was calculated by calculating the inertia values for the individual in number of subtypes [18].

K-means clustering with the initial silhouette score of values determined 2 subtypes to be the optimal model. We calculated the optimal threshold using the silhouette score of approximately

0.90. However, after assessing the predicted clusters, we determined that this model did not provide sufficient differentiation between the groups. We then attempted to use three groups classification instead which generated a more satisfactory result.

We added NT subjects to the model of classification to get a normative range of scores (as the baseline). For the validation measures, we do not have data on NT for most of them and therefore, the comparison is mainly made between subtypes.

Once the final model was selected, SPSS version 30 was used to calculate one-way ANOVAs with type III sum of squares and eta-squared effect size was used to compare subtypes and NT. Post-Hoc T-Tests with Bonferroni adjustment followed this analysis [32].

*Visualizing the Clusters of ASD*

Radar plots of the clusters of ASD comparing NEO-PI-R main and facets, converted to percentage based on min and max values within the sub-dataset and visualized created using R and the gradar package [33,34]. All other data was visualized using the Prism GraphPad software with bar plots or dot plots with standard error bars. Neuroimaging data was plotted using heatmaps of the averages for each group, with the rows being brain regions, columns being groups, and each cell the average for that group and for that individual brain region color coded as red with more connectivity and green with less connectivity [35].

*Statistical Analysis Section*

This part of the analysis was done in SPSS version 30 (IBM Corp., 2024). All data was assessed for outliers by transforming all standardized Z–Score of all values for that data and removing any data point removing individual datapoints that had a scoreless than –3 or greater than 3; the real values were for the analysis.

A one-way analysis of variance was then used to individually examine the differences in the NEO-PI-r Main and Sub Domains, RMET, BAPQ, SCL-90R, Autism Quotient, Intelligence Quotient, IRI, methylation, and resting state neuroimaging, across 4 levels including the 3 different subtypes and the NT. The neuroimaging analysis was conducted twice; once with all the subtypes and NT and again among just the subtypes. Post-Hoc T-Tests with Bonferroni adjusted P-Value were then used to examine the averages between groups.

A one-way analysis of variance was then used to individually examine the differences in the, ADOS, ADI-R, and IQ across 3 levels including the 3 different subtypes. Post-Hoc T-Tests with Bonferroni adjusted P-Value were then used to examine the averages between groups.

Restating fMRI levels of each individual brain network compared for oxytocin treatment of subjects was analyzed using a mixed model The subtypes were used as between subject's factor, while the within the four oxytocin dosages were used as the factor as a random factor while age of the subject in years an additive covariate. Follow up analysis included post-hoc T-Tests with Bonferroni adjustment and ANOVAs.

Restating fMRI levels of each individual brain network compared for oxytocin treatment of subjects was analyzed using a mixed model using SPSS version 30 [32]. The individual brain regions were used as dependent factors, with subtype and dosages and the interaction between the two as fixed factors and the subject ID as the random factor with age in years as a covariate. Type III sum of squares was used to estimates the main effects and the interaction. The variance component of the mixed model was calculated using restricted maximum likelihood. Finally, the marginal means for each different combination of subtypes and dosages were compared using a t-test with Bonferroni correction to compare differences in brain activity changes across dosages among within the individual subtypes.

For the OXTR methylation analysis, we conducted a hypothesis-based analysis focusing only on CpG 5.6 and CpG 16 sites related to the DNA methylation of *OXTR* given our previous findings (Andari et al., 2020).

Supplementary Result section

Machine learning models – Classifying ASD and NT and contributor factors

We also conducted Support vector machine analysis and found similar results with an average accuracy of 0.77 $\pm$ 0.13, an AUC of 0.84 $\pm$ 0.11, with a specificity of 0.81 $\pm$ 0.13, and with a precision of 0.84 $\pm$ 0.1, and F1 0.82 $\pm$ 0.1

Logistic Trees also had a similar score with average accuracy of 0.74 $\pm$ 0.09, an AUC of 0.81 $\pm$ 0.13, with a specificity of 0.8 $\pm$ 0.11, and with a precision of 0.81 $\pm$ 0.09, and F1 0.8 $\pm$ 0.07. Linear discriminant Analysis performed with similar accuracy, but lower AUC, score with average accuracy of 0.74 $\pm$ 0.14, an AUC of 0.75 $\pm$ 0.16, with a specificity of 0.79 $\pm$ 0.13, precision of 0.81 $\pm$ 0.11, and F1 0.8 $\pm$ 0.12. Decision Trees had the lowest accuracy of 0.7 $\pm$ 0.09, an AUC of 0.67 $\pm$ 0.11, with a specificity of 0.8 $\pm$ 0.09, precision of 0.77 $\pm$ 0.08, and F1 0.78 $\pm$ 0.07 (Figure 1A).

We found significant differences between groups in **Neuroticism** domain ($F (3, 102) = 16.508$, $P < 0.001$, eta2 = 0.327) and its 6 facets, including *Anxiety* ($F(3, 102) = 13.344$, $P < 0.001$, eta2 = 0.282), *Angry/Hostility* ($F (3, 102) = 7.827$, $P < 0.001$, eta2 = 0.187), *Depression* ($F(3, 102) = 15.742$, $P < 0.001$, eta2 = 0.316), *Self-Consciousness* ($F(3, 10) = 17.643$, $P < 0.001$, eta2 = 0.342), *Impulsiveness* ($F(3, 102) = 3.450$, $P < 0.02$, eta2 = 0.092), *Vulnerability* ($F(3, 102) = 21.321$, $P < 0.001$, eta2 = 0.385). (2A, 2E). Subtypes 1 and 3 had significantly higher scores in Neuroticism in comparison to NT ($P < 0.001$), and Subtype 2 ($P < 0.001$). Subtype 2 did not differ from NT (P = 1) but did not differ between each other (P = 1). Compared to NT and to Subtype 2, both Subtypes 1 and 3 differ in *Anxiety*, *Depression*, *Self-Consciousness*, *Vulnerability* (P < 0.005). Subtype 3 shows higher "*Angry/Hostility*" compared to Subtype 2 (P =0.041) and to NT (P < 0.001) and displays higher *Impulsiveness* compared to Subtype 2 (P =0.012) but not compared to NT (P = 0.207).

There is a significant difference between groups in **Extraversion** (F (3, 102) = 7.624, P < 0.001, eta2 = 0.183), and facets of *Warmth* (F (3, 102) = 8.275, P < 0.001, eta2 = 0.196), *Gregariousness* (F (3, 102) = 3.732, P < 0.02, eta2 = 0.099), *Assertiveness* (F (3, 102) = 4.821, P < 0.005, eta2 = 0.124), *Activity* (F (3, 102) = 4.827, P < 0.005, eta2 = 0.124), *Excitement-Seeking* (F (3, 102) = 3.34, P < 0.03, eta2 = 0.089), and *Positive Emotions* (F (3, 102) = 8.84, P < 0.001, eta2 = 0.206) (2C, 2E). Subtype 1 had significantly lower levels of Extraversion compared to Subtype 2 (P = 0.006), Subtype 3 (P = 0.012) and to NT (p < 0.001). Nothing else was significant between groups. Subtype 1 displays lower *Warmth* compared to Subtype 2, Subtype 3, and NT (P < 0.001, P = 0.013, P < 0.001), lower *Gregariousness, Assertiveness, and Positive Emotions* compared to Subtype 2 and NT (P < 0.05), and lower *Activity Level* compared to NT (P = 0.002), but no difference were found between Subtypes 2 or 3. For *Excitement Seeking*, Subtype 3 was shown to have a significantly smaller score than NT (P = 0.04).

Differences between groups were also noted for the domain of ***Openness*** *(F* (3, 102) = 7.632, P < 0.001, eta2 = 0.183), and *differences in facets of Actions (F* (3, 101) = 6.655, P < 0.001, eta2 = 0.165)*, Ideas (F* (3, 101) = 6.371, P < 0.001, eta2 = 0.159)*, and Values (F* (3, 102) = 16.824, P < 0.001, eta2 = 0.331) (2D, 2E). No differences found for *Fantasy (F* (3, 102) = 0.743, P = 0.529, eta2 = 0.021)*, Aesthetics (F* (3, 102) = 0.973, P = 0.408, eta2 = 0.028), *and Feelings (F* (3, 100) = 0.670, P = 0.573, eta2 = 0.02).

Subtype 3 showed lower scores in Openness compared to Subtype 1 and to NT (P < 0.001), but no differences were found elsewhere (P > 0.05). Subtype 3 displayed lower scores in facets of

*Values* compared to Subtype 1 and to NT (P < 0.001), in *Action* compared to NT (P < 0.001), and in *Ideas* compared to Subtype 1 and NT (P = 0.04, P < 0.001). Nothing else was significant for the other groups or other facets (2D, 2E). Subtype 2 also had lower scores in *Values* compared to Subtype 1 and NT (P = 0.001).

There is no significant difference with Agreeableness (F (3, 102) = 0.716, P = 0.545, eta2 = 0.021) nor with Conscientiousness (F (3, 101) = 0.715, P = 0.545, eta2 = 0.021).

**SCL-90-R** was shown to have significant differences with *Somatization, Obsessive-Compulsive, Interpersonal Sensitivity, Depression, Anxiety, Hostility, Phobic Anxiety, Paranoid Ideation, Psychoticism, Global Severity Index, Positive Symptom Distress Index,* and *Positive Symptom Total* F(3, 103) = 7.443, P < 0.001, eta2 = 0.178; F(3, 103) = 19.401, P < 0.001, eta2 = 0.361; F(3, 103) = 22.411, P < 0.001, eta2 = 0.395; F(3, 103) = 12.680, P < 0.001, eta2 = 0.270; F(3, 103) = 22.699, P < 0.001, eta2 = 0.398; F(3, 103) = 6.011, P < 0.001, eta2 = 0.149; F(3, 103) = 16.643, P < 0.001, eta2 = 0.326; F(3, 103) = 12.269, P < 0.001, eta2 = 0.265; F(3, 103) = 11.761, P < 0.001, eta2 = 0.255; F(3, 103) = 27.754, P < 0.001, eta2 = 0.447; F(3, 103) = 10.370, P < 0.001, eta2 = 0.232; F(3, 103) = 20.611, P < 0.001, eta2 = 0.375)

**The *intelligence quotient*** is statistically different between groups on *Verbal Comprehension Index, Perceptual Reasoning Index, Full-Scale IQ 4,* and *Full-Scale IQ 2* (F(2, 33) = 11.02, P < 0.001, eta2 = 0.4; (F(2, 33) = 6.451, P = 0.04, eta2 = 0.281; F(2, 33) = 12.807, P < 0.001, eta2 = 0.437; F(2, 33) = 10.808, P < 0.001, eta2 = 0.396).

Subtype 1 shows significantly higher scores on the *full-scale IQ-4 composite*, on the *Full-Scale IQ-2 composite*, and on the *Verbal Composite Index* in comparison to Subtypes 2 and 3 (P < 0.05). For the *Perceptual Reasoning Composite index,* Subtype 1 shows higher scores compared to Subtype 3 (P < 0.05). Subtype 2 had no difference with Subtype 3 on of the IQ sub-modules (3E).

**The *BAPQ*** sub-modules were shown to have statistically significant differences with *Aloofness, Rigidity, Pragmatic Language,* and *Average Total* (F(2, 24) = 15.582, P < 0.001, eta2 = 0.565; F(2,

24) = 5.409, P = 0.02, eta2 = 0.311; F(2, 24) = 4.671, P = 0.019, eta2 = 0.280; F(2, 24) = 11.536, P < 0.001, eta2 = 0.490) (3D).

In terms of total scores of BAPQ, Subtype 1 and Subtype 3 show higher scores compared to Subtype 2 (P < 0.005). No differences between Subtypes 1 and 3 in terms of total average scores. However, Subtype 1 does show higher sub-scores on *Aloofness* in comparison to both Subtypes 2 and 3 (P < 0.05). Subtype 1 displays higher scores in *Pragmatic Language* compared to Subtype 2 (P < 0.05), signifying more deficits in Subtype 1. Subtype 3 shows higher *Rigidity* scores on BAPQ compared to Subtype 2 (P < 0.05).

**IRI** showed significant differences in personal distress and perspective taking between 3 subtypes and NT (F (3, 43) = 11.541, P < 0.001, eta2 = 0.446; F (3, 43) = 6.453, P = 0.001, eta2 = 0.310). Post-hoc comparisons with Bonferroni corrections show that Subtype 3 shows higher personal distress compared to Subtype 2 and NT (P < 0.001). Perspective taking for Subtypes 1 and 3 is lower than NT (P < 0.05) (3F).

**Parent NEO-PI-r**, interestingly, parents' scores about their child did not yield significant differences in Neuroticism between subtypes which is contrary to the self-report of Neuroticism dimension of the personality test (F (2, 27) = 0.252, P = 0.779). Parents' scores on ASD individuals did show differences between subtypes for Extraversion (F (2, 27) = 3.588, P = 0.042) and for Openness (F (2, 27) = 7.749, P< 0.005). Post-hoc tests with Bonferroni corrections show that

Subtype 1 had higher scores on Openness, values and ideas than Subtypes 2 and 3 (P < 0.05).

Subtype 3 shows lower positive emotions compared to Subtype 2 (P < 0.005) (S3).

Therefore, if Subtype 3 is perceived by others as less positive or less cheerful, Subtype 1 shows high self-report of neuroticism and self-report of less positive emotions and extraversion is not perceived as negative by their surroundings. Subtype 3 also displays high self-report of neuroticism and is not perceived as such by their parents.

**Clinical validation:**

ADI-R with clinician yielded significant differences between subtypes for the ADI repetitive behaviors and restricted interests ($F(2,30) = 5.941$, $P = 0.007$, $eta2 = 0.284$). No differences were found for the other sub scores (social, communication) or total score ($P > 0.05$). Subtype 3 has significantly higher ADI-R repetitive behaviors than Subtypes 1 and 2 ($P < 0.05$) (S4).

No significant differences were found for the ADOS-2 severity calibration scores.

**Biological validation of the 3 putative subtypes:**

a- Resting-state functional connectivity results:

- For the 3 subtypes and NT, connectivity between several networks revealed significant differences between groups using ANOVA analysis and post-hoc t-tests with Bonferroni corrections (See Figure 5A, 5B):
  - Sensorimotor network to theory of mind network (STS) ($F(3, 41) = 4.289$, $P = 0.010$, $eta2 = 0.239$). Post-hoc comparisons revealed that Subtype 1 has significantly lower rsFC between these networks compared to NT ($P = 0.021$).
  - Theory of Mind (STS) to Visual network ($F(3, 41) = 3.582$, $P = 0.022$, $eta2 = 0.208$). Subtype 3 shows significant hypoconnectivity compared to NT ($P<0.05$).
  - Empathy/salience network (ACC, AI) and Visual cortex ($F(3, 40) = 3.442$, $P = 0.026$, $eta2 = 0.205$). Subtype 3 shows hypoconnectivity compared to NT ($P<0.05$).
  - Somatosensory network to Brainstem/cerebellum network ($F(3,41) = 4.4104$, $P = 0.012$, $eta2 = 0.231$). Subtype 3 shows hyperconnectivity within between these networks compared to NT ($P<0.05$).

  - Ventromedial PFC (or default network) and sensorimotor network ($F(3,41) = 3.086$, $P = 0.038$, eta2 = 0.184). However none of the post-hoc tests were significant.
  - Ventromedial PFC (or default network) and Fusiform Occipital network ($F(3, 41) = 2.969$, $P = 0.043$, eta2 = 0.178). However none of the post-hoc tests were significant.
- We then compared 3 ASD subtypes between each other using exploratory t-tests and we found the following results:
  - Sensorimotor network to theory of mind network (STS) differences between Subtype 1 had lower connectivity then subtype 2 ($t(14) = -2.375$, $P = 0.032$, Cohen's $d = -1.197$).
  - Default mode (posterior cingulate cortex) and visual cortex showing Subtype 1 have lower connectivity than Subtype 2 ($t(14) = -3.720$, $P = 0.002$; Cohen's $d = -1.875$).
  - Reward network to dorsolateral prefrontal areas ($t(14) = -2.296$, $P = 0.038$, Cohen’s $d = -1.157$) showing lower connectivity in Subtype 1 compared to Subtype 2.
  - Somatosensory to DL-PFC ( $t(14) = -2.558$, $P = 0.023$, Cohen’s $d = -1.289$) showing lower connectivity in Subtype 1 compared to Subtype 2.
  - Default mode network (vmPFC) and inferior frontal gyrus showing higher connectivity for Subtype 3 compared to Subtype 1 ($t(16) = -2.266$, $P = 0.038$, $d = -1.096$) and Subtype 2($t(18) = -2.608$, $P = 0.009$, $d = -1.172$).
  - Subtype 3 shows also higher rsFC between default mode network (vmPFC) and fusiform/occipital areas compared to subtype 2 ($t(19) = -2.232$, $P = 0.038$, $d =-0.984$).

b- DNA methylation of the oxytocin receptor gene:

- We conducted a hypothesis-based analysis focusing only on CpG 5.6 and CpG 16 sites related to the DNA methylation of *OXTR* given our previous findings (Andari et al., 2020). Subtype 3 had higher *OXTR* DNA methylation at CpG 5.6 site compared to Subtype 1 ($t(22) = -1.953$, $P = 0.032$ with one-sided p value, Cohen's $D = -0.809$). No differences were found between Subtypes 1 and 2 ($t(16) = -0.802$, $P = 0.217$, Cohen's $D = -0.381$) nor between Subtypes 2 and 3 ($t(20) = -0.885$, $P = 0.193$, Cohen's $D = -0.392$). No differences were found for CpG 16 site between subtypes (5C, 5D).

**2.4 Personalized drug treatments – Intranasal oxytocin (Syntocinon spray Novartis) as one potential drug treatment.**

We used mixed models that include different doses of IN-OXT as a random factor, resting-state functional networks as dependent factor, subtypes a fixed factor, and age as a covariate to examine if there is a particular subtype that is more impacted by IN-OXT than others.

As mentioned in the methods section, these data are part of a clinical trial (NCT03033784; https://clinicaltrials.gov/study/NCT03033784?cond=autism&term=andari&rank=1) that was performed on ASD adults with different doses of IN-OXT (8IU, 24IU, 48IU) and intranasal placebo in a randomized fashion. Data includes resting-state functional networks effects which were part of a previous publication for a different purpose (Andari et al., 2025) to study effects of drug on rsFC. Here, we are using the data to classify ASD into different groups and to see if there is a subtype that has more modulatory effects from IN-OXT.

We investigated whether there is interaction between the different factors (subtypes of ASD, all doses of IN-OXT, and rsFC networks) and found the following results (see table 1):

Cerebellum to PCC Default (2to20): There was a statistically significant interaction between subtype and dosage F (6, 74.107) = 3.850, P = 0.002. For subtype 1, 48 IU (M = 0.415, SE = 0.085) had significant increase compared to placebo (M = 0.108, SE = 0.085) (P = 0.007). 8 IU for Subtype 1 (M = 0.409, SE = 0.085) had significantly higher brain activity then placebo (P = 0.009). Subtype 3 with 48 IU (M = 0.419, SE = 0.064) had increased brain activity when compared to 24 IU. (M = 0.176, SE = 0.064) and 8 IU (M = 0.195, SE = 0.064) (P = 0.005, P = 0.011).

Cerebellum Brainstem to Parietal Lobe (1to12): There was a statistically significant interaction between subtype and dosage F (6, 75.039) = 2.437, P = 0.03. For Subtype 1, 48 IU (M = -0.173, SE = 0.071) had significantly decreased compared to 24 IU (M = 0.077, SE = 0.071) and placebo (M = 0.095, SE = 0.071) (P =0.038, P = 0.021).

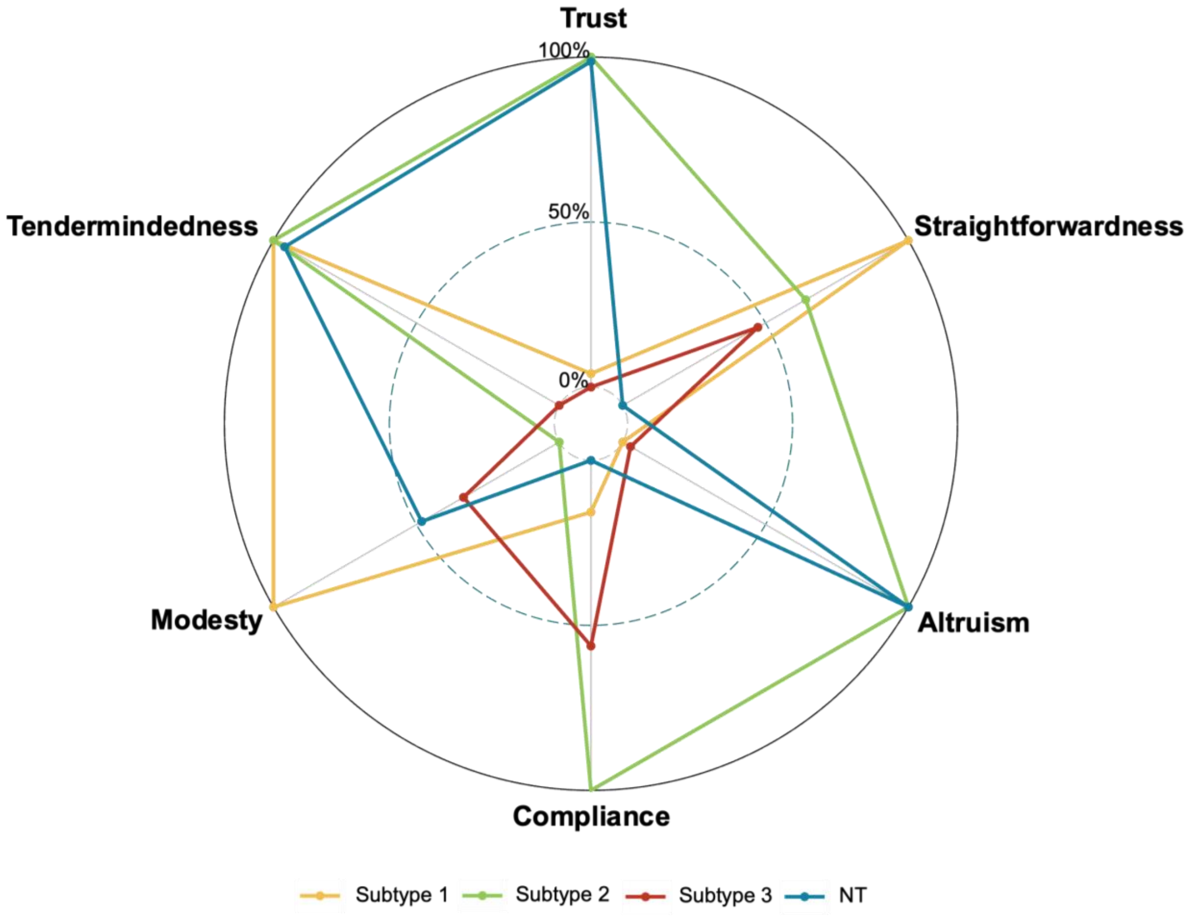


**Figure S1: Agreeableness Radar Plot**

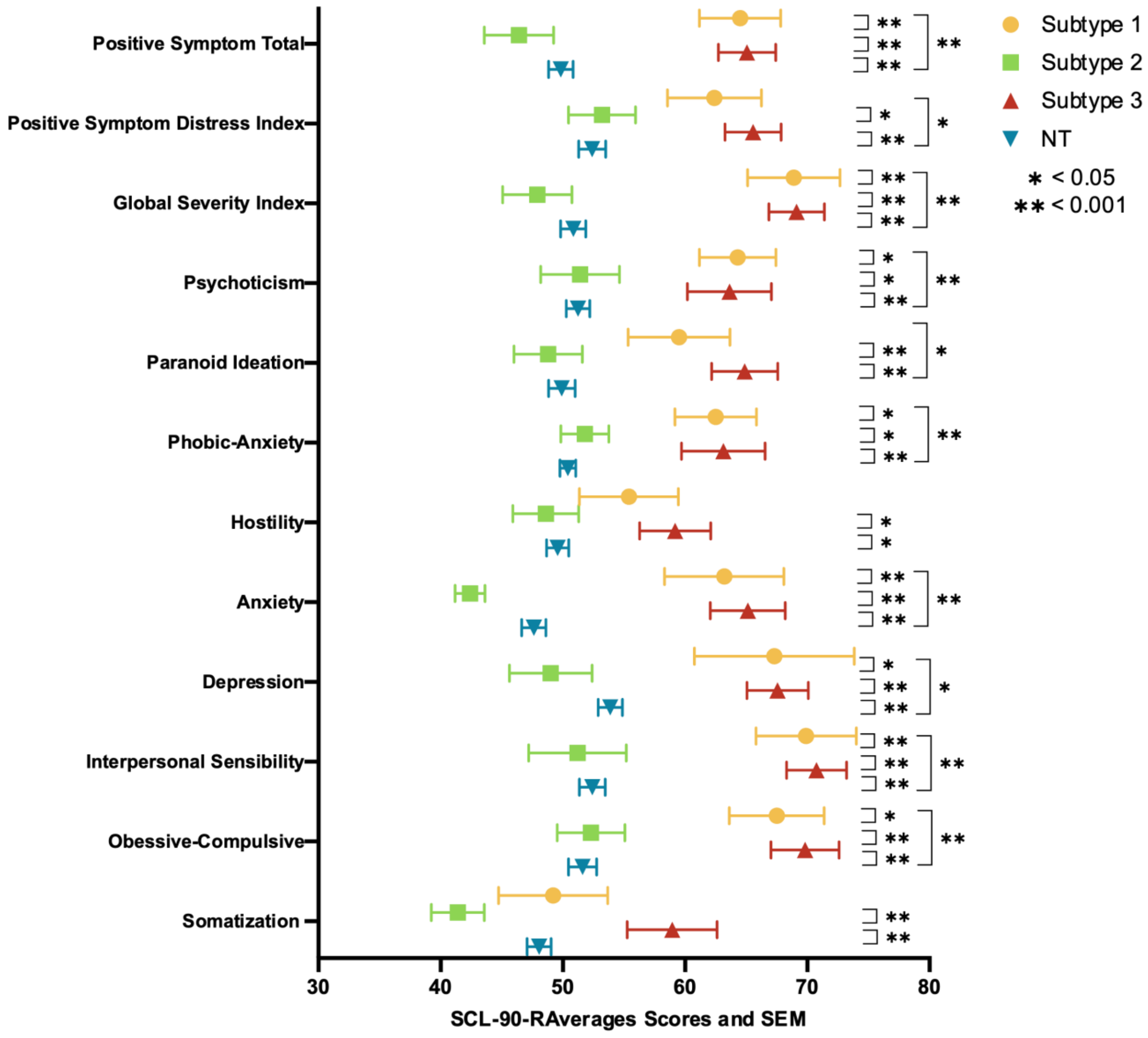


**Figure S2: SCL-90-R Average Scores**

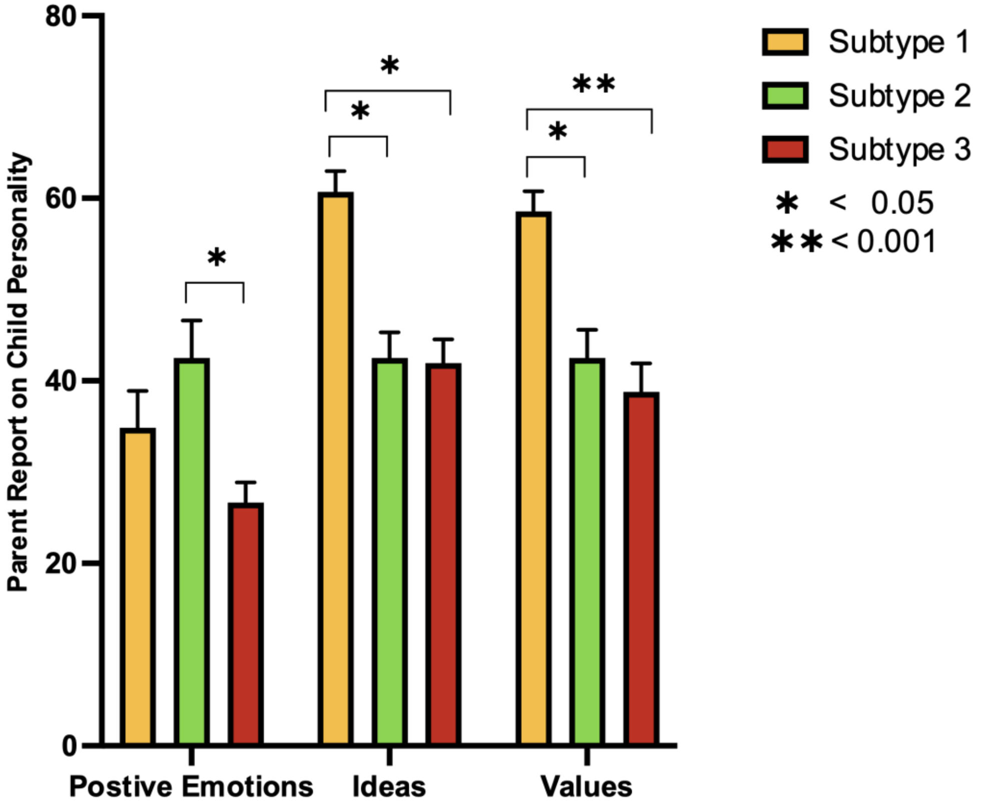


**S3: Parent NEO-PI-R for Positive Emotions, Ideas, and Values**

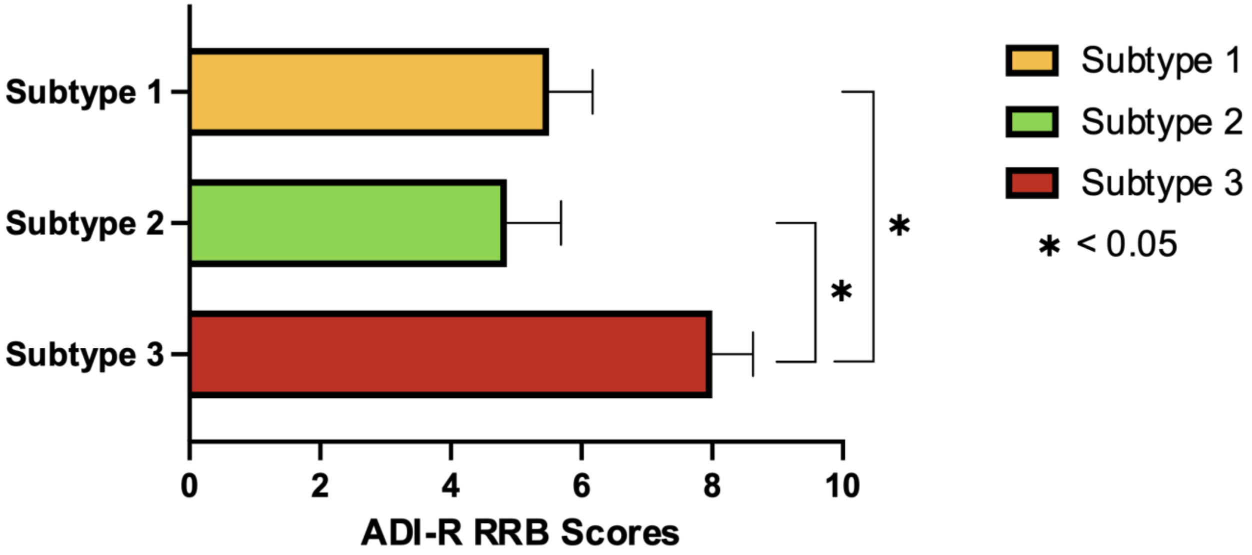


**S4: ADI-R Repetitive Behavior Scores**

22. Kusui C, Kimura T, Ogita K, Nakamura H, Matsumura Y, Koyama M, et al. DNA Methylation of the Human Oxytocin Receptor Gene Promoter Regulates Tissue-Specific Gene Suppression. Biochem Biophys Res Commun. 2001;289:681–686.
23. Kogan SM, Cho J, Beach SRH, Smith AK, Nishitani S. Oxytocin receptor gene methylation and substance use problems among young African American men. Drug Alcohol Depend. 2018;192:309–315.
24. Smith SM, Jenkinson M, Woolrich MW, Beckmann CF, Behrens TEJ, Johansen-Berg H, et al. Advances in functional and structural MR image analysis and implementation as FSL. NeuroImage. 2004;23:S208–S219.
25. Calhoun VD, Liu J, Adali T. A review of group ICA for fMRI data and ICA for joint inference of imaging, genetic, and ERP data. NeuroImage. 2009;45:S163-172.
26. Greve DN, Fischl B. Accurate and robust brain image alignment using boundary-based registration. NeuroImage. 2009;48:63–72.
27. Pruim RHR, Mennes M, van Rooij D, Llera A, Buitelaar JK, Beckmann CF. ICA-AROMA: A robust ICA-based strategy for removing motion artifacts from fMRI data. NeuroImage. 2015;112:267–277.
28. Allen EA, Erhardt EB, Damaraju E, Gruner W, Segall JM, Silva RF, et al. A Baseline for the Multivariate Comparison of Resting-State Networks. Front Syst Neurosci. 2011;5.
29. Yarkoni T, Poldrack RA, Nichols TE, Van Essen DC, Wager TD. Large-scale automated synthesis of human functional neuroimaging data. Nat Methods. 2011;8:665–670.
30. Smith SM, Fox PT, Miller KL, Glahn DC, Fox PM, Mackay CE, et al. Correspondence of the brain's functional architecture during activation and rest. Proc Natl Acad Sci. 2009;106:13040–13045.
31. Van Rossum G, Drake FL. Python 3 Reference Manual. Scotts Valley, CA: CreateSpace; 2009.
32. IBM Corp. IBM SPSS Statistics for Windows (Version 30.0). 2024.
33. R Core Team. R: A Language and Environment for Statistical Computing. Vienna, Austria: R Foundation for Statistical Computing; 2023.
34. Bion R. ggradar: Create radar charts using ggplot2. 2023.
35. GraphPad Prisim version 10.0.0 for Windows.
